# A novel multi-omics mendelian randomization method for gene set enrichment and its application to psychiatric disorders

**DOI:** 10.1101/2024.04.14.24305811

**Authors:** Huseyin Gedik, Roseann Peterson, Christos Chatzinakos, Mikhail G. Dozmorov, Vladimir Vladimirov, Brien P. Riley, Silviu-Alin Bacanu

**Affiliations:** Integrative Life Sciences, Virginia Institute of Psychiatric and Behavioral Genetics, Virginia Commonwealth University, Richmond, VA, USA; Institute for Genomics in Health, SUNY Downstate Health Sciences University, Brooklyn, NY, USA; Department of Psychiatry and Behavioral Sciences, SUNY Downstate Health Sciences University, Brooklyn, NY, USA; Department of Psychiatry, McLean Hospital, Harvard Medical School, Belmont, MA, USA; Stanley Center for Psychiatric Research, Broad Institute of MIT and Harvard, Cambridge, MA, USA; Department of Biostatistics, Virginia Commonwealth University, Richmond, VA, USA; Department of Psychiatry, College of Medicine-Phoenix, University of Arizona, Phoenix, AZ, USA; Department of Psychiatry, Virginia Institute for Psychiatric and Behavioral Genetics, Virginia Commonwealth University, Richmond, VA, USA

**Keywords:** pQTL, eQTL, mQTL, GSMR, schizophrenia, AMPA

## Abstract

Genome-wide association studies (GWAS) of psychiatric disorders (PD) yield numerous loci with significant signals, but often do not implicate specific genes. Because GWAS risk loci are enriched in expression/protein/methylation quantitative loci (e/p/mQTL, hereafter xQTL), transcriptome/proteome/methylome-wide association studies (T/P/MWAS, hereafter XWAS) that integrate xQTL and GWAS information, can link GWAS signals to effects on specific genes. To further increase detection power, gene signals are aggregated within relevant gene sets (GS) by performing gene set enrichment (GSE) analyses.

Often GSE methods test for enrichment of “signal” genes in curated GS while overlooking their linkage disequilibrium (LD) structure, allowing for the possibility of increased false positive rates. Moreover, no GSE tool uses xQTL information to perform mendelian randomization (MR) analysis. To make causal inference on association between PD and GS, we develop a novel MR GSE (MR-GSE) procedure. First, we generate a “synthetic” GWAS for each MSigDB GS by aggregating summary statistics for x-level (mRNA, protein or DNA methylation (DNAm) levels) from the largest xQTL studies available) of genes in a GS. Second, we use synthetic GS GWAS as exposure in a generalized summary-data-based-MR analysis of complex trait outcomes.

We applied MR-GSE to GWAS of nine important PD. When applied to the underpowered opioid use disorder GWAS, none of the four analyses yielded any signals, which suggests a good control of false positive rates. For other PD, MR-GSE greatly increased the detection of GO terms signals (2,594) when compared to the commonly used (non-MR) GSE method (286). Some of the findings might be easier to adapt for treatment, e.g., our analyses suggest modest positive effects for supplementation with certain vitamins and/or omega-3 for schizophrenia, bipolar and major depression disorder patients. Similar to other MR methods, when applying MR-GSE researchers should be mindful of the confounding effects of horizontal pleiotropy on statistical inference.

## Introduction

Psychiatric disorders (PD) are debilitating illnesses with complex etiology (Uher and Zwicker, 2017). Heritability of PD varies from 40-80% depending on the specific condition; genetic risk factors are thus an important component of PD risk (Sullivan et al., 2012). Large scale genome wide association studies (GWAS), relying on both measured and imputed SNPs, are currently widely used in genetic studies. One of the key benefits of using post-imputation data from the PGC is the vast increase in SNP coverage, which allows for the detection of some associations that might be missed by employing only directly genotyped data (Sullivan et al., 2018). [However, uncertainty in imputed genotypes can introduce noise in analysis and may affect the accuracy of association signals (Pasaniuc and Price, 2017).] The use of this post-imputation GWAS paradigm assisted researchers in detecting numerous loci with significant association signals (Wray et al., 2018; Mullins et al., 2021; Trubetskoy et al., 2022). However, its usage rarely identified specific risk loci that can elucidate the molecular underpinnings of PD. One of the reasons is that detected GWAS signal is just in linkage disequilibrium (LD) with the causal genetic marker (Visscher et al., 2012). Second, most GWAS associated loci are not in protein coding regions (Maurano et al., 2012), but signals can be driven by intergenic enhancers (Ernst et al., 2011) or long-range acting (Schierding et al., 2014; Mifsud et al., 2015)] and intronic (Smemo et al., 2014; Claussnitzer et al., 2015) regulatory sequence elements. Consequently, it is challenging to translate these discoveries into viable drug targets and therapeutic approaches for PD. To add biological meaning to GWAS findings, researchers developed post-GWAS methods that aggregate GWAS signals at gene and gene set (GS) level.

One approach to aggregate variant information at gene level entails the use of molecular mediator data obtained from large-scale QTL mapping studies, e.g., expression quantitative trait loci (eQTL) (Qi et al., 2018; Võsa et al., 2021), protein QTL (pQTL) (Ferkingstad et al., 2021; Wingo et al., 2021) and DNA methylation (DNAm) QTL (mQTL) (McRae et al., 2018; Qi et al., 2018; Wu et al., 2018) studies. These reference e/p/mQTL (hereafter, xQTL) datasets provide empirical information regarding the effect of genetic variants on the level of gene expression, protein or DNAm (hereafter, x-level). Subsequently, transcriptome, proteome and methylome wide association studies (TWAS, PWAS or MWAS, hereafter XWAS) methods integrate the reference xQTL with GWAS summary statistics to infer the association statistics between the x-level (gene expression/protein/DNAm level) and complex disorders, such as PD (Li et al., 2021; Wingo et al., 2021). Mendelian randomization (MR) approaches (Davey Smith and Ebrahim, 2003; Lawlor et al., 2008) offer alternative ways to perform XWAS by testing for more causally-oriented inferences for the effect of x-level on trait (Zhu et al., 2016; Yuan et al., 2020; Zhou et al., 2020). One such method is the summary-data-based mendelian randomization (SMR) that implements a MR gene-level test (Zhu et al., 2016) using only summary statistics for both GWAS and x-level.

Most xQTL studies to date have been performed using expression, protein or DNAm levels from bulk tissue. For disorders of the central nervous system like PD, this is a limitation because of the complex mix of cell types in brain, with cell type specific active regulatory elements and transcriptome profiles specific to their functions. It is important to consider cell type specificity of xQTL and single cell eQTL reference datasets that are becoming available, although they are still limited for brain cell types. Functional genomic profiles are different among these diverse cell types (Marstrand and Storey, 2014; Buenrostro et al., 2015) and the cellular make-up shows variation throughout various areas of brain (Wang et al., 2018). Association signals for genetic variants that are linked with distinct PD were identified to be enriched in cis-sequence elements with regulatory function that are exclusively found in neurons responsible for GABA and glutamate signaling (Sanchez-Priego et al., 2022). For instance, when the genetic risk for schizophrenia (SCZ) was attributed to different types of neurons (cortical pyramidal neurons and cortical interneurons) (Skene et al., 2018). Consequently, the integration of cell type-specific xQTL data with genome-wide association study (GWAS) results holds significant potential. However, due to financial and other constraints, cell type specific functional profiling across brain regions has still limitations in terms of sample sizes (Spaethling et al., 2017; Bryois et al., 2022) and this results in lowered power to detect signals. Although the cell-level resolution is somewhat restricted, meta-analyzing multiple tissues and using bulk tissue are the still viable ways to have more detection power. Hence, in this study we will mainly focus on inferences regarding blood and brain XWAS statistics. Moreover, while mQTL and other QTL might be of interest to many researchers, we focused solely on eQTL and pQTL reference studies because of i) their larger sample sizes and ii) the direct link between such xQTL and genes.

While XWAS aggregates the variant information at the gene level and improves the detection power, this may not always be enough to completely elucidate the molecular basis of disorders. To further improve detection power and replicability of findings, researchers proposed aggregating information at the GS level (Li et al., 2011). These methods are denoted as gene set enrichment (GSE) and they utilize lists of relevant GS that are available from various reference databases, such as BioCarta (Nishimura, 2001), Gene Ontology (GO) (The Gene Ontology Consortium, 2021), Kyoto Encyclopedia of Genes and Genomes (KEGG) (Kanehisa et al., 2008, 2017), Molecular Signatures Database (MSigDB) (Subramanian et al., 2005; Liberzon et al., 2011), Panther (Mi et al., 2013), Pathway Commons (Cerami et al., 2011), Reactome (Jassal et al., 2020) and Wiki Pathways (Martens et al., 2021). Such information is, subsequently, used by GSE methods for making statistical inferences, like the first such method: gene set enrichment analysis (Subramanian et al., 2005; Watanabe et al., 2017). Also, researchers proposed other methods, such as the database for annotation, visualization and integrated discovery (DAVID) (Dennis et al., 2003; Huang et al., 2009; Sherman et al., 2022), gene-set enrichment analysis (Wang et al., 2007), g:Profiler (Reimand et al., 2007), correlation adjusted mean rank gene set test (Camera) (Wu and Smyth, 2012), interval-based enrichment analysis (Lee et al., 2012), data-driven expression prioritized integration for complex traits (Pers et al., 2015), multi-marker analysis of genomic annotation (MAGMA) (Leeuw et al., 2015), pathway scoring algorithm (Lamparter et al., 2016) and functional mapping and annotation (FUMA) (Watanabe et al., 2017).

Many of the above-mentioned methods use hypergeometric test to obtain the enrichment statistics, which ignores the possible LD structure, and none of them attempts to perform any type of causal inference. Because some MR methods, e.g., generalized summary-data-based MR (GSMR), provide a more causal inference alongside a heterogeneity in dependent instruments (HEIDI) outlier test (Zhu et al., 2018), which eliminates the effect of the local LD structure. It would be advantageous to apply these to be used for causal inference at GS level. Unfortunately, while numerous MR methods exist for gene level inference, to our best knowledge, there is no available MR-type GSE method yet. Consequently, developing such a tool would fill the methodological gap – especially if its usage requires only summary statistics for trait GWAS, e.g., like GSMR. We note that such a GS MR inference can be easily performed if a relevant “synthetic” GS GWAS can be computed (in silico) and, subsequently, used as exposure in a GSMR analysis of trait GWAS.

In this study, we developed a Mendelian randomization GSE (MR-GSE) method. We achieved this by computing a synthetic GS GWAS by combining for each SNP, via a Simes test (Li et al., 2011), the x-level GWAS statistics for all genes in the investigated GS. We applied MR-GSE to GWAS of nine PD using synthetic GWAS summary statistics generated by aggregating (for both brain and blood) eQTL/pQTL statistics for MSigDB GS. Applying MR-GSE to the underpowered opioid use disorder GWAS produced no significant signal in any of the four different analyses, which strongly suggests that the proposed method effectively controls the false positive rates. Signals for the better powered disorders tend to fall mainly into synapse/neuronal, immune, cytoskeleton, RNA translation, and chromatin accessibility, miRNA, apoptosis and “housekeeping” categories. However, there are also a few unexpected findings.

## Methods

To uncover risk GS for complex disorders, we proposed an innovative method termed Mendelian randomization gene set enrichment (MR-GSE) that makes a causal inference on GS using an integrative analysis based on GSMR (Zhu et al., 2018). One important advantage of using GSMR is that it incorporates a multivariate method (HEIDI-outlier test) to identify and eliminate variants that are involved in horizontal pleiotropy (i.e., they influence exposure and trait independently), which can confound other MR estimates. By eliminating pleiotropic variants, and thus horizontal pleiotropy, GSMR ensures the validity of the critical MR assumption that SNP, serving as instrumental variables, act on the outcome solely via their effects on the exposure.

To perform MR-GSE, the main idea is to first aggregate x-level GWAS summary statistics, obtained from the largest available eQTL/pQTL meta-analyses for blood/brain, into MSigDB GS summary statistics using MSigDB collection. These statistics were subsequently used as exposure to identify putatively causal GS associated with PD (Figure 1). (Next paragraph provides a more detailed description of this process.) By using all x-level GWAS summary statistics available, our MR method ensures the inclusion of both cis- and trans-xQTL. Recognizing that genes within GS might not affect the trait in the same direction, the MR-GSE tests whether departures from mean x-level are associated with departures from mean risk for disease. In practice, the departure from mean x-level/risk is achieved by using an upper quantile gaussian transformation for the p-values of both exposure and trait GWAS.

MR-GSE has two major steps (Figure 1): the construction of synthetic GWAS for GS [i.e., curated GS in MySigDB (v2022.1Hs) (Liberzon et al., 2011)] and using them, alongside trait GWAS, in a GSMR inference. To obtain the synthetic GWAS of a GS, we first aggregate, for each SNP, p-values for genes in GS into a single q-value by applying Simes test (Simes, 1986). In more detail, the Simes test is applied to the vector consisting of p-values for the SNP in x-level GWAS associated with all genes in investigated GS. Second, this q-value serves as the SNP p-value in the synthetic GWAS, which is subsequently converted to an upper tail SNP Z-scores to account for the above-mentioned unknown direction of effect on risk. Third, we calculate SNP slope effect in synthetic GS GWAS as the product of its Z-score and the average of the standard errors of SNP slopes from x-level GWAS associated with each gene in GS. (See Supplementary Material section 3 for more details on increasing computational efficiency for generating synthetic GS GWAS.) The last two steps from computing GS GWAS were also applied to trait GWAS, albeit using SNP p-value in the second step and standard error of SNP’s slope estimate in the third step. Next, we used GS and trait GWAS to compute MR-GSE test by conducting a two sample GSMR analysis to estimate the effects of GS (exposure) on the PD (outcome).

**Figure 1.**
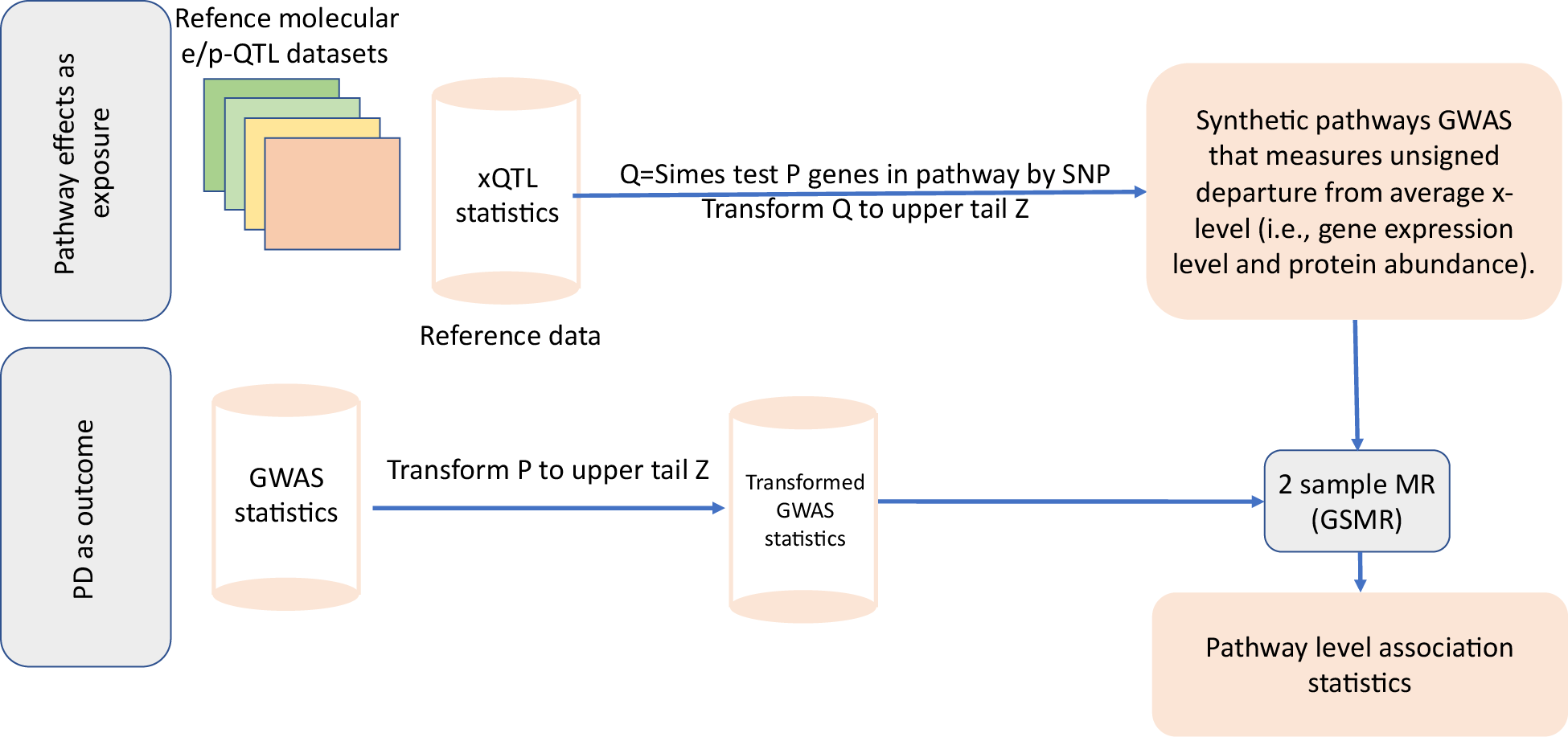
Overall workflow for MR-GSE analysis using two sample MR (GSMR).

In practical terms, we computed XWAS GS statistics by using GSMR option from Genome-wide Complex Trait Analysis (v1.94.1) (Yang et al., 2011) and the LD reference panel from 1000 Genomes phase 3 including only European ancestry (Auton et al., 2015). GS were included in the GSMR analysis only if their associated synthetic GWAS had at least 10 “independent” xQTL SNP with p-value < 5×10^-8^ (< 10^-6^ for brain pQTL since power of this study was lower). As suggested by authors of other GS methods (e.g., GSEA), we initially filtered out MSigDB GS with too few (<10) and too many (>500) genes. When generating synthetic GS GWAS, we further excluded GS if they had <5 genes with available x-level GWAS. (This last filtering step was necessary because, for instance, brain pQTL reference studies had assayed only a few thousand proteins.)

To also use the HEIDI outlier information from GSMR alongside SMR effect size, we derived a new (adjusted) p-value for each XWAS GS test by penalizing its original value (*P_GSMR_*) for low HEIDI p-values: 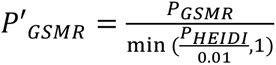. We opted for this p-value adjustment, rather than using a *P_HEIDI_* cutoff < 0.01. because it is always possible that there is a mismatch in ancestry structure between the LD reference panel and the GWAS cohort in GSMR analysis. Such a mismatch might result in significantly low *P_HEIDI_* values (as encountered by our group when performing a gene level SCZ TWAS analysis of *C4A*, which yielded a *P_HEIDI_* < 1×10^-4^ alongside a much smaller *P_GSMR_*).

To control for multiple testing, within each paradigm (blood and brain for two xQTL types) we used the Benjamini & Hochberg False Discovery Rate (FDR) procedure (Benjamini and Hochberg, 1995) to obtain the q-value that was associated with each GS. We deemed a GS test as significant (or a signal) if FDR < 0.05 (FDR – referred hereafter simply as q-value).

### Investigated psychiatric disorders

We applied MR-GSE to uncover GS signals for nine major PD (Table 1). For eight of them, we obtained summary statistics for their latest publicly available GWAS, the Psychiatric Genomics Consortium (PGC) (https://pgc.unc.edu/for-researchers/download-results/, accessed on 12/7/2022): autism spectrum disorder (ASD), attention deficit and hyperactivity disorder (ADHD), cannabis use disorder (CUD), bipolar disorder (BIP), major depression (MDD), opioid use disorder (OUD), post-traumatic stress disorder (PTSD) and schizophrenia (SCZ). For the ninth disorder, alcohol use disorder (AUD), we obtained summary statistics from Million Veteran Program (MVP) (Kranzler et al., 2019) via dbGAP (accession number: phs001672.v6.p1). (See Supplementary material for details).

**Table 1.** Cohort information for GWAS of nine major PD.

| Psychiatric disorders | Study | Number of cases | Number of controls | Continental ancestry |
| --- | --- | --- | --- | --- |
| ADHD | (Demontis et al., 2019) | 19,099 | 34,194 | EUR |
| ASD | (Grove et al., 2019) | 18,381 | 27,969 | EUR |
| **AUD** | (Kranzler et al., 2019) | 34,658 | 167,346 | EUR |
| **BIP** | (Mullins et al., 2021) | 41,917 | 371,549* | EUR |
| **CUD** | (Johnson et al., 2020) | 20,196 | 363,116 | EUR |
| **MDD** | (Howard et al., 2019) | 411,965 | 1,285,068 | EUR |
| **OD** | (Polimanti et al., 2020) | 4,503 | 32,500 | EUR |
| **PTSD** | (Nievergelt et al., 2019) | 30,000 | 170,000 | EUR |
| **SCZ** | (Trubetskoy et al., 2022) | 33,640 | 43,456 | EUR** |

EUR: European
\* Cases were subjects with bipolar or unipolar. Controls did not have any of these two disorders.
\*\* predominantly European

### Transcriptome and proteome reference data sets

To maximize detection power for XWAS GS analyses, we sourced the x-level GWAS from the largest xQTL reference datasets that were publicly available, i.e., the latest brain and blood meta-analyses (studies) (Ferkingstad et al., 2021; Võsa et al., 2021; Wingo et al., 2021; Qi et al., 2022) (see also online resources for more details). Briefly, we downloaded blood eQTL summary statistics (n = 31,684) from eQTLGen consortium (Võsa et al., 2021), which assayed the collected samples via gene expression array and RNA sequencing platforms. Brain eQTL data (n = 2,865) were obtained from a meta-analysis (Qi et al., 2018, 2022) of several different studies (BrainGVEX, Common Mind Consortium, Lieber Institute for Brain Development, Mayo Clinic, Mount Sinai VA Medical Center Brain Bank, NIMH Human Brain Collection Core, Religious Order Study and Rush Memory and Aging). We downloaded blood pQTL data (n = 35,559) from a large-scale study by deCODE (Ferkingstad et al., 2021) that was assayed using SOMAScan (Gold et al., 2010, 2012). Brain pQTL data were obtained from a recent study (Wingo et al., 2022), for which subjects (n = 722) were assayed using mass spectrometry-based method (Johnson et al., 2018).

### Gene set data

Our analyses are based on gene sets coming from MSigDB (v2022.1.Hs; accessed on January 19 2023, http://www.gsea-msigdb.org/gsea/downloads.jsp). It is a well-known resource that built its collection based on input from expert opinion or solid empirical research. While we did not include the entire MSigDB collection, we selected those gene sets coming from expert opinion (GO, BIOCARTA, KEGG, WP) or likely to be relevant to our research: curated gene sets collection – C2 (n = 6,448), regulatory target gene sets – C3 (predicted as miRNA or transcription factor binding) (n = 3,725), ontology gene sets – C5 (n = 15,703; excluded human phenotype ontology), oncogenic signature gene sets – C6 (n = 189) and immunologic signature gene sets – C7 (n = 5,219; excluded vaccination response gene sets, VAX). Further details on these collections can be found in Supplementary Table S1.

### Competing method

For comparison with existing approaches, we also obtained gene-level XWAS statistics, and for the more permissive “suggestive” signals (HEIDI adjusted *p′_SMR_* < 1/number of tests), performed a GS enrichment (Gedik et al., 2023). The XWAS GS enrichment of these signals was performed using FUMA – gene-to-function tool (Watanabe et al., 2017). We computed the FUMA enrichments statistics using the same MSigDB collection that was used to generate MR-GSE results (Supplementary Table S1).

## Results

MR-GSE analyses yielded n = 8,193 signals with FDR < 0.05 across all disorders (except OUD) in both brain (n = 6,695) and blood (n = 1,498) (Table 2). FUMA analysis, even using suggestive instead of significant signals, yielded only 1,407 signals. MR-GSE consistently outperformed FUMA except for blood TWAS (for BIP, MDD, PTSD and SCZ) and blood PWAS (for ASD, AUD and CUD). The better performance of FUMA for blood TWAS, was likely due to only cis-eQTL information being publicly available for download (in addition to the use of the more permissive suggestive signals for FUMA).

**Table 2.**
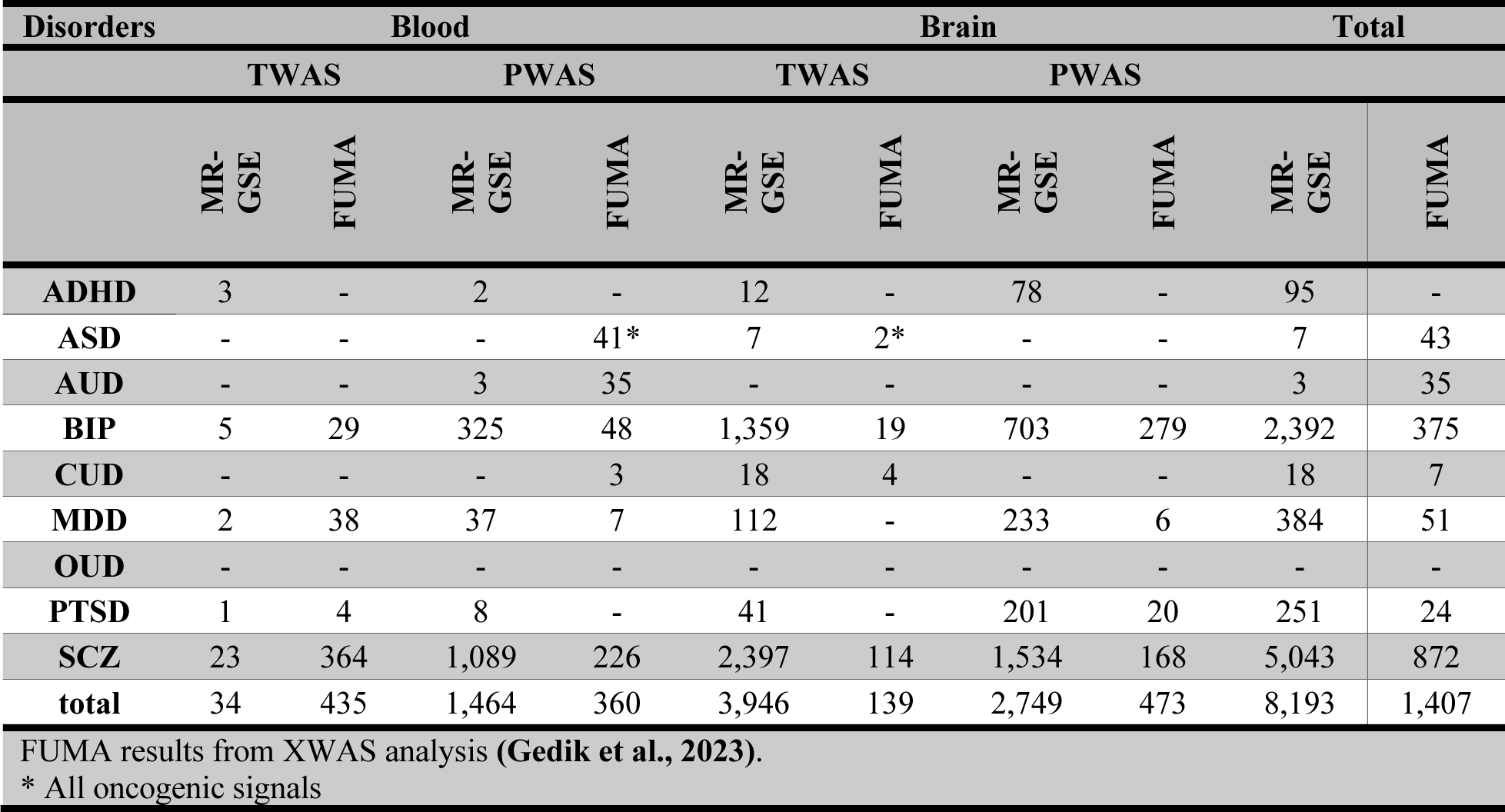
Signal distribution by disorder and tissue.

| Disorders | Blood |  |  |  | Brain |  |  |  | Total |  |
| --- | --- | --- | --- | --- | --- | --- | --- | --- | --- | --- |
|  | TWAS |  | PWAS |  | TWAS |  | PWAS |  | MR-GSE | FUMA |
|  | MR-GSE | FUMA | MR-GSE | FUMA | MR-GSE | FUMA | MR-GSE | FUMA |  |  |
| ADHD | 3 | - | 2 | - | 12 | - | 78 | - | 95 | - |
| ASD | - | - | - | 41* | 7 | 2* | - | - | 7 | 43 |
| AUD | - | - | 3 | 35 | - | - | - | - | 3 | 35 |
| BIP | 5 | 29 | 325 | 48 | 1,359 | 19 | 703 | 279 | 2,392 | 375 |
| CUD | - | - | - | 3 | 18 | 4 | - | - | 18 | 7 |
| MDD | 2 | 38 | 37 | 7 | 112 | - | 233 | 6 | 384 | 51 |
| OUD | - | - | - | - | - | - | - | - | - | - |
| PTSD | 1 | 4 | 8 | - | 41 | - | 201 | 20 | 251 | 24 |
| SCZ | 23 | 364 | 1,089 | 226 | 2,397 | 114 | 1,534 | 168 | 5,043 | 872 |
| total | 34 | 435 | 1,464 | 360 | 3,946 | 139 | 2,749 | 473 | 8,193 | 1,407 |
| FUMA results from XWAS analysis (Gedik et al., 2023). |  |  |  |  |  |  |  |  |  |  |
| * All oncogenic signals |  |  |  |  |  |  |  |  |  |  |

The number of significant GS signals observed among PD varies considerably (BIP and SCZ having highest number of signals across tissues and TWAS/PWAS), which is related to the power of GWAS – as measured by the number of GWAS signals (Supplementary Figure S1). Consequently, the lack of GS signals detected for OUD could be due to limitations in the sample size of cohorts with European ancestry. (On the other hand, the underpowered OUD not yielding any significant signal in any of the four analyses suggests that MR-GSE controls false positive rates.) In more details, SCZ yielded the largest number of significant signals (n = 5,043), followed by BIP (n = 2,392), MDD (n = 384), PTSD (n = 251), ADHD (n = 95), CUD (n = 18), ASD (n=7) and AUD (n = 3) (Table 2). ASD signals being all oncogenic corroborates previous findings that show autism and cancer are negatively correlated (Liu et al., 2022).

When compared to blood, brain analyses yielded a higher number of signals for most disorders, except for AUD – where three signals occurred in blood. The lowest number of blood GS signals was for ASD, CUD and OUD which do not have any signals (n=0). Besides the underpowered OUD, there were no brain signals for AUD. The PD with highest number of blood (n=1,112; 74.23%) and brain (n=3,931; 58.72%) signals was SCZ. Thus, while the brain generally exhibited a higher number of signals, blood might still provide valuable information about PD e.g., biomarkers for PD.

We stress that many of these signals are not independent due to their overlapping genes across GS. Consequently, we present the results by broader and biologically relevant GS categories (e.g., immune, neuronal, apoptosis, cytoskeleton etc.), which are less likely to have a high degree of overlap between their GS. Detailed results can be found in Supplementary Tables S2-S5 (also at https://huseyingedik.shinyapps.io/mrgse/, accessed on Dec 1 2023).

### TWAS GS analysis results

The sequence for presenting MR-GSE findings consists of first overviewing GS results for each individual PD and then providing details on shared signals among PD. There were fewer signals in ADHD, ASD, CUD and AUD when compared to other PD (Table 2). The strongest signal for ADHD was ACOSTA PROLIFERATION INDEPENDENT MYC TARGETS UP (q-value = 1.15×10^-2^) in brain. We also found unexpected ADHD blood signals for heart morphogenesis, such as GOBP ATRIAL SEPTUM MORPHOGENESIS (q-value = 1.24×10^-2^) and GOBP MITRAL VALVE DEVELOPMENT (q-value = 1.24×10^-2^), which might point to the existence of common developmental pathway for ADHD and some heart conditions. For ASD, the strongest signal was apoptosis related (REACTOME DISEASES OF PROGRAMMED CELL DEATH – q-value = 1.25×10^-3^) (Supplementary Table S6). Among the disorders with fewer signals, CUD yielded two strong brain signals (also shared with SCZ, see Figure 2) for a neuronal (GOBP POSITIVE REGULATION OF CALCIUM ION DEPENDENT EXOCYTOSIS – q-value = 1.66×10^-6^), immune (GOMF TAP BINDING – q-value = 2.94×10^-4^) and an unexpected (GOBP NEGATIVE REGULATION OF REPRODUCTIVE PROCESS – q-value = 1.66×10^-6^) GS (Supplementary Table S6). (Related reproductive process GS also yielded weaker signals for BIP and SCZ).

**Figure 2.**
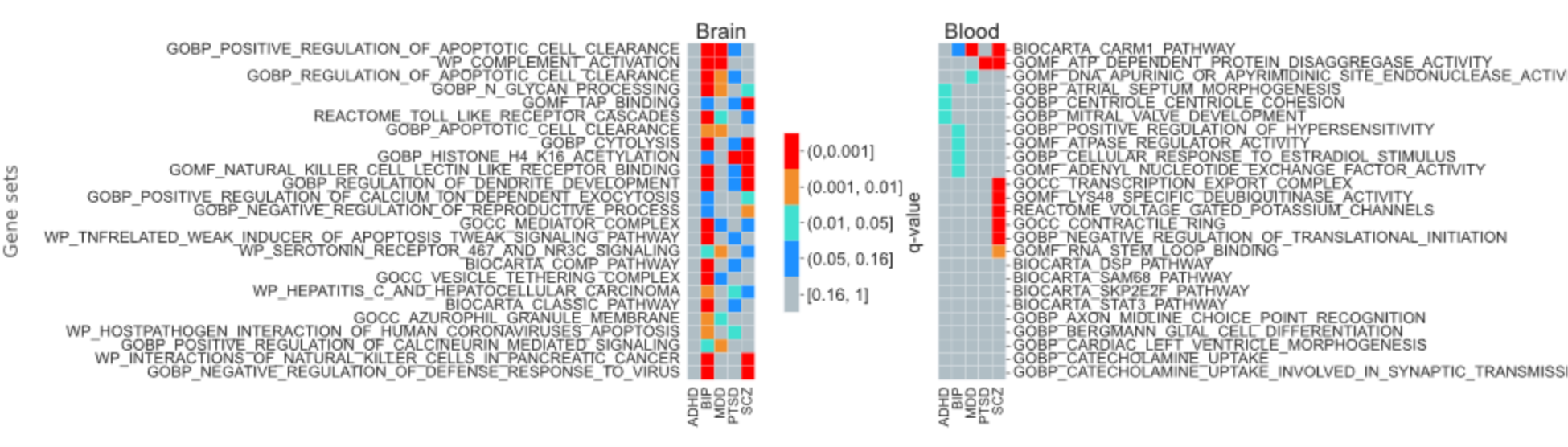
TWAS GS results for five PD with most signals. Color denotes FDR q-value intervals as detailed in legend. To underscore shared GS, signals were ranked by the sum (across all disorders) of categorical values (between 1 and 5) corresponding, in ascending order, to the above five p-value intervals. For conciseness, we included only GS from GO, BIOCARTA, REACTOME and WikiPathways (WP).

For BIP, several miRNA GS yielded stronger significant GS signals, e.g., MIR140_3P (q-value = 8.44×10^-10^), which was the strongest signal in the brain. Notable wiki pathway (WP) GS signals in brain TWAS were WP COMPLEMENT ACTIVATION (q-value = 6.71×10^-5^), WP NEUROINFLAMMATION AND GLUTAMATERGIC SIGNALING (q-value = 5.17×10^-4^), WP CANNABINOID RECEPTOR SIGNALING (q-value = 7.01×10^-3^), vitamin metabolism (WP VITAMIN A AND CAROTENOID METABOLISM – q-value = 4.24×10^-2^), WP SEROTONIN RECEPTOR 467 AND NR3C SIGNALING (q-value = 2.41×10^-2^) and WP COMPLEMENT ACTIVATION (q-value = 6.71×10^-5^).

MDD yielded brain signals for some neuron (GOCC INTRINSIC COMPONENT OF POSTSYNAPTIC DENSITY MEMBRANE – q-value = 9.93×10^-3^) and serotonin (WP SEROTONIN RECEPTOR 467 AND NR3C SIGNALING – q-value = 3.52×10^-2^) related GS. MIR6776_3P (q-value = 4.20×10^-6^) was the strongest association for MDD in brain. This miRNA has an overlapping 17p13.3 microdeletion associated with SCZ (Warnica et al., 2015). In addition to this, several apoptotic related GS were detected in brain, such as GOBP POSITIVE REGULATION OF APOPTOTIC CELL CLEARANCE (q-value = 9.73×10^-4^) and GOBP INTRINSIC APOPTOTIC SIGNALING PATHWAY IN RESPONSE TO DNA DAMAGE (q-value = 7.62×10^-3^). A notable cluster of GS in brain was immune related GS, e.g., WP COMPLEMENT ACTIVATION (q-value = 2.78×10^-5^) and GOBP TUMOR NECROSIS FACTOR SUPERFAMILY CYTOKINE PRODUCTION (q-value = 1.14×10^-3^).

For PTSD, notable signals were detected for histone modification (GOBP HISTONE H4 K16 ACETYLATION – q-value=1.02×10^-7^), immune (GOCC MHC CLASS I PROTEIN COMPLEX – q-value=1.22×10^-2^) and miRNA (MIR1914_5P – q-value = 1.50×10^-2^) related GS. Expression of MIR1914_5P in brain has been suggested to be unique to synapse in individuals with a closely related disorder (MDD) (Yoshino et al., 2021).

For SCZ, there were brain signals for neuronal GS, e.g., WP DISRUPTION OF POSTSYNAPTIC SIGNALING BY CNV (q-value = 6.31×10^-11^) and GOBP SYNAPSE ORGANIZATION (q-value = 1.45×10^-7^). Another possibly biologically relevant SCZ brain signal was for WP SEROTONIN HTR1 GROUP AND FOS PATHWAY (q-value = 1.12×10^-8^). The validity of the serotonin system’s role in SCZ treatment and pathology has been intensely debated over time (Tauscher et al., 2002). Another interesting SCZ brain signal occurred for a drug addiction (WP COMMON PATHWAYS UNDERLYING DRUG ADDICTION – q-value = 8.55×10^-11^), which seem to support previous findings that SCZ shares genetic factors with substance use disorders (Jang et al., 2022). Other notable brain related GS were GOBP REGULATION OF POSTSYNAPTIC MEMBRANE NEUROTRANSMITTER RECEPTOR LEVELS (q-value = 1.28×10^-6^) and GOBP DENDRITE MORPHOGENESIS (q-value = 3.39×10^-6^). Also, a number of miRNA target GS (MIR382_5P, MIR4430 and MIR3652) yielded strong signals. Previous research findings suggest an increased expression of MIR382_5P in cells derived from patients diagnosed with SCZ (Mor et al., 2013).

To identify GS that are most likely to be shared between disorders we i) excluded underpowered disorders (ASD, AUD, CUD and OUD) and ii) ranked GS signals by the sum over disorders of their FDR q-value “intensity bin” with values 1-5, which correspond to the following five q-value intervals: (0.16, 1], (0.05, 0.16], (0.01, 0.05], (0.001, 0.01] and < 0.001 (see also Figure 2 legend). For brain, GS signals fall into six main categories (Figure 2). (Please see Supplementary Table S6 for all significant GS with FDR < 0.05; also, at https://huseyingedik.shinyapps.io/mrgse/, accessed on May 16 2023) First one was apoptotic/lysosomal GS (e.g., GOBP POSITIVE REGULATION OF APOPTOTIC CELL CLEARANCE/GOBP N-GLYCAN PROCESSING) that were shared especially between BIP and MDD (Figure 2). The second category was immune-related (e.g., WP COMPLEMENT ACTIVATION, REACTOME TOLL LIKE RECEPTOR CASCADES), which yielded significant signals for BIP and MDD. Third was histone modification (chromatin accessibility - we used these two terms interchangeably) (e.g., HISTONE H4 K16 ACETYLATION) that was shared between PTSD and SCZ. This histone modification was associated with HOX9 gene expression which is involved in embryonic development (Dou et al., 2005)

The fourth category was neuronal (GOBP REGULATION OF DENDRITE DEVELOPMENT and GOBP REGULATION OF DENDRITE MORPHOGENESIS) GS, which were shared between BIP and SCZ. Fifth was “housekeeping” (e.g., GOBP CYTOLYSIS) that largely assures the well-functioning of the cell. [The housekeeping GS were classified based on the list of genes from recent research findings (there should be more than 50% of genes from this list in GS to be considered as housekeeping) (Funk et al., 2022)] Sixth category was miRNA. While these do not occur in Figure 2 there are 738 miRNA related GS (Supplementary Table S6). Some of these miRNA signals were shared between more than two disorders: BIP, PTSD and SCZ (MIR1914_5P and MIR6836_3P) as well as BIP, MDD and SCZ (MIR218_5P and MIR6856). Numerous miRNA signals are shared between two disorders: BIP and SCZ (e.g., MIR382_5P, MIR140_3P and MIR4430) as well as MDD and SCZ (such as MIR6776_3P).

In blood, there were fewer shared signals than in brain. The strongest shared signal was for RNA translation/chromatin accessibility (BIOCARTA CARM1 PATHWAY), which was shared between MDD and SCZ (Figure 2). The second strongest signal was for a housekeeping GS (GOMF ATP DEPENDENT PROTEIN DISAGGREGASE ACTIVITY), which yielded signal of PTSD and SCZ. Notably, there were no miRNA signals in blood TWAS analyses.

There are also some unexpected, albeit modest, signals for vitamins and omega-3 GS. BIP yielded brain signals for REACTOME METABOLISM OF FAT SOLUBLE VITAMINS (q-value = 2.38×10^-2^), an empirical vitamin D response GS (GSE13762 CTRL VS 125 VITAMIND DAY12 DC UP – q-value = 4.26×10^-2^) and WP VITAMIN A AND CAROTENOID METABOLISM (q-value = 4.24×10^-2^) and SCZ just for the later (q-value = 3.98×10^-2^). SCZ also yielded a brain signal for WP OMEGA3 OMEGA6 FATTY ACID SYNTHESIS (q-value = 3.93×10^-2^).

### PWAS GS analysis results

There were no signals detected in ASD, CUD and OUD and fewer signals in ADHD when compared to other PD (Table 2). The strongest GS signals for ADHD were cytoskeleton related ones detected in brain PWAS, such as, GOCC MICROTUBULE and GOCC SPINDLE. Among the more underpowered traits, we mention the biologically relevant signals for AUD in blood for GOBP ETHANOL OXIDATION (q-value = 2.79×10^-2^) (Supplementary Table S6).

In BIP, the strongest signal was GOMF TRANSLATION FACTOR ACTIVITY RNA BINDING which is translation related GS. In addition to this, there were several other translations related GS signals were detected in blood, e.g., WP TRANSLATION FACTORS and BIOCARTA EIF PATHWAY. Several miRNA target GS also yielded significant signal: MIR6779_5P, MIR30B_3P, MIR8078 and MIR5002_3P.

The strongest signal for MDD was in brain and it was immune related (GSE19512 NAUTRAL VS INDUCED TREG_UP – q-value = 2.70×10^-8^). There were several other strong signals for immune related differential expression GS, such as GSE26928 CENTR MEMORY VS CXCR5 POS CD4 TCELL UP (q-value = 9.33×10^-8^) and GSE13229 MATURE VS INTMATURE NKCELL UP (q-value = 2.01×10^-7^), which corroborate the focus on the role of immune systema and inflammation in MDD etiology (Ruiz et al., 2022). We also detected significant signals for synapse related GS, e.g., GOCC PRESYNAPTIC ACTIVE ZONE (q-value = 3.90×10^-5^) and GOBP SYNAPTIC VESICLE RECYCLING (q-value = 5.56×10^-4^). Recent findings suggest a strong association at pathway level association between synapse biology and MDD (Fries et al., 2023). Another group of GS was apoptosis and cell death, such as GOBP REGULATION OF CELL KILLING (q-value = 7.47×10^-3^) and REACTOME FORMATION OF APOPTOSOME (q-value = 3.29×10^-2^). For miRNA target GSE12003 MIR223KO VS WT BM PROGENITOR 8D CULTURE DN yielded a strong signal, which corroborates the empirical association between MIR223 (Zhao et al., 2019) and SCZ in plasma of patients at first episode. Another miRNA GS signal was for MIR5002_3P, which was also previously associated with SCZ (O’Connell et al., 2019).

The three strongest signals for PTSD were GOBP REGULATION OF CHRONIC INFLAMMATORY RESPONSE (blood; q-value = 3.80×10^-10^), GOBP POSITIVE REGULATION OF TYPE 2 IMMUNE RESPONSE (blood; q-value = 4.66×10^-10^) and GOBP CHRONIC INFLAMMATORY RESPONSE (blood; q-value = 6.10×10^-7^). When compared with all PTSD TWAS signals, there were relatively higher number of immune related experimental differential expression GS signals were also detected in brain, e.g., GSE3039 ALPHAALPHA CD8 TCELL VS B2 BCELL UP (q-value = 2.37×10^-5^). Cytoskeleton (GOCC ACTIN BASED CELL PROJECTION – q-value = 2.28×10^-3^) and brain (GOBP NEURON PROJECTION GUIDANCE – q-value = 2.37×10^-5^) related signals were significant in brain.

The strongest SCZ signal was for glutamate receptor (REACTOME TRAFFICKING OF GLUR2 CONTAINING AMPA RECEPTORS – q-value = 4.45×10^-37^). We detected synapse and neuron related GS, such as GOBP NEGATIVE REGULATION OF NERVOUS SYSTEM_DEVELOPMENT (q-value = 2.74×10^-7^), GOCC GLUTAMATERGIC SYNAPSE (q-value = 2.48×10^-4^), GOBP NEURON PROJECTION ORGANIZATION (q-value = 3.66×10^-4^) and GOBP VESICLE MEDIATED TRANSPORT IN SYNAPSE (q-value = 2.94×10^-3^). A majority of significant signals in brain belong to a cluster of immune system related GS, such as GSE27786 LIN NEG VS BCELL UP (q-value = 1.72×10^-4^) and GOBP POSITIVE REGULATION OF LYMPHOCYTE MEDIATED IMMUNITY (q-value = 5.08×10^-9^). Another important cluster of signals were detected for cytoskeleton and cell-cell adhesion molecules GS, e.g., GOBP CYTOSKELETON DEPENDENT INTRACELLULAR TRANSPORT (q-value = 9.54×10^-5^) and GOBP REGULATION OF CELL CELL ADHESION (q-value = 3.34×10^-4^). Possible involvement of cell adhesion molecules corroborates recent findings on association between cerebral levels of these molecules and SCZ risk (Sheikh et al., 2023). An unexpected finding in blood was WP BILE ACIDS SYNTHESIS AND ENTEROHEPATIC CIRCULATION (q-value = 3.86×10^-22^) and GOBP REGULATION OF BILE ACID METABOLIC PROCESS (q-value = 1.01×10^-11^).

However, recent empirical research suggests an association between level of bile acid in serum and SCZ risk (Qing et al., 2022). In blood, MIR8078 (q-value = 3.83×10^-31^), MIR5002_3P (q-value = 5.96×10^-27^) and MIR4468 (q-value = 2.26×10^-22^) were strongest miRNA-related signals.

While decidedly more modest than signals in other categories, there are reasonably strong findings for some vitamin related GS. For instance, SCZ yielded a brain signal for GOBP WATER SOLUBLE VITAMIN METABOLIC PROCESS (q-value = 2.37×10^-4^), and a blood signal for GOBP REGULATION OF VITAMIN METABOLIC PROCESS (q-value = 3.80×10^-2^) GS. BIP and MDD had a signal for an empirical vitamin D response GS (GSE13762 CTRL VS 125 VITAMIND DAY12 DC UP – q-value = 4.26×10^-2^; GSE13762 CTRL VS 125 VITAMIND DAY5 DC UP – q-value = 1.22×10^-3^).

There were also shared PWAS GS signals in the brain (Figure 3). These signals fall largely into six categories. First, immune (e.g., GOBP REGULATION OF IMMUNE EFFECTOR PROCESS) GS yielded signals for BIP, MDD and SCZ. Second, neuronal (e.g., GOBP VESICLE MEDIATED TRANSPORT IN SYNAPSE, GOBP NEURON PROJECTION) GS were shared between BIP and at least one of MDD, SCZ and PTSD. In addition to these, GOBP SYNAPTIC VESICLE RECYCLING yielded brain signals for BIP, MDD and SCZ. Third, cytoskeleton related GS (e.g., GOBP CYTOSKELETON DEPENDENT CYTOKINESIS) were significant signals for BIP, PTSD and SCZ. Fourth, adhesion (GOMF CADHERIN BINDING) GS yielded common significant signals for BIP, MDD and SCZ. Fifth, some housekeeping (e.g., GOBP REGULATION OF NUCLEOTIDE METABOLIC PROCESS, GOBP REGULATION OF OXIDOREDUCTASE ACTIVITY) GS were also shared by BIP, MDD and SCZ. Sixth, empirical miRNA GS were shared between BIP, MDD and SCZ (MIR5002_3P and MIR6890_5P), MDD and SCZ (MIR173P and MIR595) and, BIP and SCZ (MIR223 and MIR155_5P) (Supplementary Table S6).

**Figure 3.**
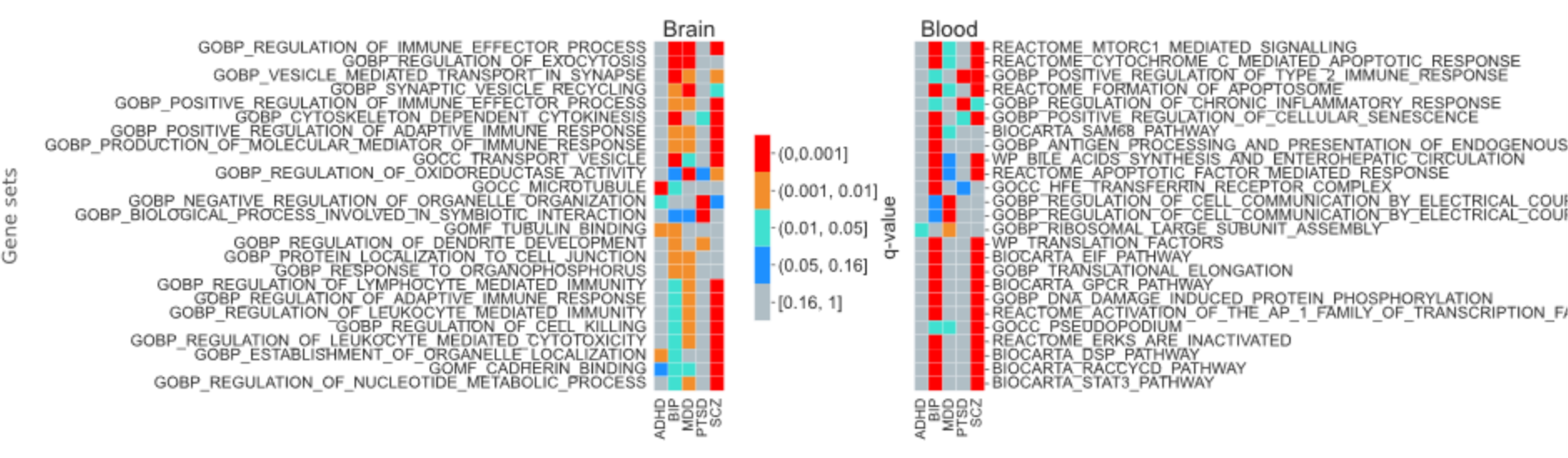
PWAS GS results for the five disorders with most signals.

In blood, shared signals fall into six categories (Figure 3). First, immune related (e.g., GOBP POSITIVE REGULATION OF TYPE 2 IMMUNE RESPONSE) GS yielded signal for BIP, PTSD and SCZ. In the same category, REACTOME AMINO ACIDS REGULATING MTORC1 was shared between BIP, MDD and SCZ. Second, RNA translation (e.g., WP TRANSLATION FACTORS, BIOCARTA EIF PATHWAY and GOBP TRANSLATIONAL ELONGATION) GS yielded signals for BIP and SCZ. Third, lysosome/apoptosis related (e.g., REACTOME CYTOCHROME C MEDIATED APOPTOTIC RESPONSE and REACTOME FORMATION OF APOPTOSOME) GS were shared between BIP, MDD and SCZ. Fourth, housekeeping (e.g., BIOCARTA GPCR PATHWAY) GS yielded signals for BIP and SCZ. Fifth, it is worth highlighting unexpected GS relating to bile acid circulation (WP BILE ACIDS SYNTHESIS AND ENTEROHEPATIC CIRCULATION) and synthesis (GOBP REGULATION OF BILE ACID METABOLIC PROCESS) that yielded signals for BIP and SCZ. Sixth, miRNA, e.g. the strong shared signals for BIP and SCZ (MIR8078, MIR6779_5P, MIR30B_3P and MIR3689A_3P_MIR3689B_3P_MIR3689C), BIP, MDD and SCZ (MIR5002_3P) and, MDD and SCZ (MIR595) (Supplementary Table S6).

While not quite among the strongest shared signals, GS related to the glutamatergic system also exhibited significant brain signals for BIP and SCZ. For example, GOCC GLUTAMATERGIC SYNAPSE was shared between BIP (q-value = 2.01 x 10^-3^) and SCZ (q-value = 2.48 x 10^-4^). Same two disorders also shared REACTOME ACTIVATION OF KAINATE RECEPTORS UPON GLUTAMATE BINDING in brain TWAS (BIP) and blood PWAS (SCZ) (q-values = 4.08×10^-3^ and 1.89×10^-2^, respectively). In the same vein, the most significant GS for SCZ in blood was also the glutamate related REACTOME TRAFFICKING OF GLUR2 CONTAINING AMPA RECEPTORS (q-value = 4.45×10^-37^).

## Discussion

The goal of this study was to develop a novel MR approach for GS enrichment analysis of complex disorders and use it to uncover putative causal risk GS for PD. Previously, such an MR approach did not exist due to the lack of a GS level GWAS that can serve as the exposure in a GSMR analysis. We addressed this shortcoming by developing MR-GSE, which generates *in silico* GWAS for each MSigDB GS based on x-level (xQTL) summary statistics of all genes within that GS. We applied our method to nine PD using the largest publicly available xQTL reference data for blood and brain. In terms of number of detected signals, MR-GSE greatly outperformed FUMA, even though FUMA used the more permissive XWAS suggestive signals (Table 2). However, it underperformed FUMA for blood TWAS, where only cis-eQTL information was available. MR-GSE controlled the false positive rates and identified several “positive controls”, e.g., the alcohol metabolism GS for AUD.

Neuron related GS were some of the common XWAS signals (Figures 2 and 3). The strongest signal for SCZ was REACTOME TRAFFICKING OF AMPA RECEPTORS. Although this signal was found in blood, there were brain signals in related GS (GOCC GLUTAMATERGIC SYNAPSE and GOCC AMPA GLUTAMATE RECEPTOR COMPLEX) for the same disorder (Supplementary Table S7). Previously, loci with GWAS signals were found to be enriched glutamate receptor genes in the PGC cross disorder study (Lee et al., 2019). There were also several significant BIP signals for GS in SynGO (Koopmans et al., 2019) (Supplementary Table S8). Synaptic GS signals were previously detected in BIP GWAS (Mullins et al., 2021), although it was not based on XWAS results. Corroborating the serotonin theory of MDD, there were signals for this disorder in WP SEROTONIN RECEPTOR 467 AND NR3C SIGNALING. In parallel with this, the same signal occurred for BIP, which confirms a recent BIP finding for a serotonin related GS (Cabana-Domínguez et al., 2022). Also, WP SEROTONIN HTR1 GROUP AND FOS PATHWAY yielded a SCZ brain TWAS signal (Supplementary Table S7), which supports the finding that serotonin receptor and transporter genes were associated with SCZ (Hrovatin et al., 2020).

Immune related GS yielded numerous signals for i) BIP, MDD and SCZ (GOBP REGULATION OF IMMUNE EFFECTOR PROCESS) in brain PWAS (Figure 3 and Supplementary Table S6), ii) BIP, MDD and (WP COMPLEMENT ACTIVATION) in brain TWAS (Figure 2) and iii) BIP and SCZ (GOBP HUMORAL IMMUNE RESPONSE) in brain TWAS (Supplementary Table S6). In addition, GOBP COMPLEMENT ACTIVATION (also a signal for SCZ) and REACTOME INITIAL TRIGGERING OF COMPLEMENT were significant for BIP in brain TWAS. While it was not fully understood how the immune system takes part in the pathology of PD, *C4* (complement component 4), in the MHC region, was shown to be involved in synaptic pruning in a model organism (Sekar et al., 2016).

RNA translation and histone modification GS form another important category of signals. WP TRANSLATION FACTORS and GOBP TRANSLATIONAL ELONGATION were shared blood signals between BIP and SCZ (Figure 3). Previously, cells derived from patients with SCZ showed elevated levels of mRNA for translation complex related proteins, such as initiation and elongation factors (Topol et al., 2015). Histone modification GS yielded significant brain signals relating to histone methylation (GOBP POSITIVE REGULATION OF HISTONE METHYLATION and GOBP POSITIVE REGULATION OF HISTONE H3 K4 METHYLATION for SCZ, GOBP REGULATION OF HISTONE H3 K4 METHYLATION for BIP), histone acetylation (GOBP HISTONE H4 K16 ACETYLATION for SCZ and PTSD, GOBP HISTONE H4 ACETYLATION for BIP), acetyltransferase complex (GOCC H4 HISTONE ACETYLTRANSFERASE COMPLEX for BIP) and deacetylase complex (GOMF HISTONE DEACETYLASE BINDING for MDD and SCZ). Histone modification signatures could affect the pathogenesis of PD. For instance, previous findings found an association between H3K4 trimethylation level in promoter regions of synapsin genes (which have a role in synaptogenesis) and differential expression of these genes in postmortem brains from individuals with BIP and MDD (Cruceanu et al., 2013). Also, SCZ patient derived neuron cells as well as cells in post-mortem tissues presented hypermethylation in H2A.Z and H4 (Farrelly et al., 2022).

MicroRNAs and post-transcriptional gene regulation were hypothesized to be involved in PD etiology (Geaghan and Cairns, 2015) and our results support this assertion, e.g., some of the strongest BIP and SCZ signals were for miRNA target GS (Supplementary Table S6). While we mostly discussed BIP and SCZ miRNA findings, we note that MIR6776_3P targets yielded the ninth strongest brain TWAS MDD signal and ASD seventh strongest signal was for MIR3059_3P targets in brain TWAS. Some of these miRNA signals were shared between more than two disorders: BIP, PTSD and SCZ (MIR1914_5P and MIR6836_3P)

For BIP, the two strongest miRNA targets [MIR6779_5P (q-value = 9.15×10^-27^) and MIR30B_3P (q-value = 4.22×10^-26^)] were also overlapping with the following GS in MSigDB: GOCC SYNAPSE and GOCC NEURON PROJECTION. For SCZ, MIR5002_3P targets a set of genes that overlaps with GOBP CELLULAR RESPONSE TO MANGANESE ION (q-value = 2.76×10^-3^), which was also significant in blood PWAS for BIP and SCZ (Supplementary Table S6). Manganese (Mn) is known as a trace element that, in excess amounts, becomes neurotoxic (Stredrick et al., 2004) and could interfere with synaptic transmission (Takeda, 2003). miR-5002 has been implicated in SCZ for its possible regulatory role on *CACNA1B* (O’Connell et al., 2019).

MIR140_3P GS was detected in brain TWAS for both BIP and SCZ. This miRNA showed a differential expression in the postmortem prefrontal cortex from individuals with BIP (Kim et al., 2010). Other examples of our miRNA GS signals that were previously implicated in differential expression and functional studies are: MIR382 (Santarelli et al., 2011) and MIR223 (Zhao et al., 2019) in SCZ, and MIR4430 (Yoshino et al., 2020) in MDD. Previous association studies suggest MIR6836 as risk loci in dimensional psychopathology (McCoy et al., 2018) and MIR6776 was implicated in copy number variant in SCZ (Warnica et al., 2015). We also detected miRNA GS signals, which were previously pinpointed in GWAS (albeit not using MR inferences): MIR137 for SCZ (Ripke et al., 2011), MIR5007 for PTSD (Nievergelt et al., 2019) and MIR2113 for BIP (Mühleisen et al., 2014)

Apoptosis related processes are conspicuously involved in many PD, e.g., apoptosis/lysosomal (e.g., GOBP POSITIVE REGULATION OF APOPTOTIC CELL CLEARANCE/GOBP N-GLYCAN PROCESSING) GS were shared significant signals in brain TWAS between BIP, MDD and (sometimes) SCZ (Figure 2). Similarly, even the more underpowered ASD yielded its strongest signal for REACTOME PROGRAMMED CELL DEATH in brain TWAS (Supplementary Table S6). Apoptosis is known to be involved in brain development (Kuan et al., 2000). Post-mortem brain samples of children with ASD had a rise in neuron count in prefrontal cortex (Courchesne et al., 2011). It is likely that apoptosis impairment leads to a dysfunctional control for the number of neurons, which, in turn, may lead to this abnormality in a number of neurons for individuals with ASD.

ADHD yielded signals for several cytoskeleton-related GS (e.g., microtubule organization) (Supplementary Tables S6 and S9), which sometimes also yielded signals for BIP and SCZ. These findings corroborate previous research that suggested the involvement of the cytoskeleton in ADHD pathology and, specifically, due to its possible role in neurite growth (Poelmans et al., 2011). In addition to this, the loci with CNV that are associated with ADHD were enriched in cytoskeleton GS (Harich et al., 2020). Empirical research suggested the existence of differential case-control methylation of *MARK2* in ADHD (Mooney et al., 2020), which takes part in microtubule organization (Drewes et al., 1997) and dendrite growth (Biernat et al., 2002).

However, there were also some unexpected findings. For instance, signals in heart morphogenesis GS were also detected for ADHD in blood TWAS (Figure 2). Based on health records, the adults with ADHD showed an increased risk for cardiovascular diseases (Li et al., 2022). A study of a large health registry also suggested an increased risk for cardiovascular disease in individuals with ADHD (Rietz et al., 2021). Unfortunately, it was not clear from previous epidemiological research whether cardiac abnormalities are due to common causes with ADHD or due to the medication used to treat it. However, our results suggest the existence of shared genetic risk factors between the two disorders. In blood PWAS, BIP and SCZ yielded unexpected signals for bile acid synthesis and circulation. Previous research showed differences in bile acid composition between SCZ patients and the control group (Tao et al., 2021; Qing et al., 2022).

While exhibiting more modest signals, certain vitamin and omega-3 GS yielded mostly brain signals, with the strongest one coming from brain PWAS. This suggests the investigation of supplementing (as complementary) drug treatment for SCZ with water soluble vitamins and omega-3. For BIP and MDD, our results suggest that vitamin D supplementation can also be investigated further when its concentration in the blood is low. However, as these signals were generally modest, such supplementation for PD is likely to have a modest effect overall, likely being useful only in subjects with very low biological reserves for such supplements.

This study is important for the field for four main reasons. First, it proposes MR-GSE that is a novel and powerful MR-based GS method. Second, the proposed method can use both cis- and trans-xQTL due to its whole genome aggregation of SNP summary statistics from x-level GWAS for genes in each MSigDB GS. Third, the PD MR-GSE analyses presented in this study were based on the largest publicly available xQTL reference datasets. Fourth, the synthetic GS GWAS is available to assist researchers in studying other phenotypes.

## Limitations

The xQTL reference datasets were mostly obtained from assayed pre-frontal cortex. So, it might not be relevant for substance use disorders that have been associated with cerebellum in addition to other brain regions (Carta et al., 2019). Also, the brain pQTL reference dataset was not as extensive as the other xQTL data, which diminishes its detection power. Trans-xQTL require more sample size for discovery power, consequently, GS signals can be tested for sensitivity by performing similar analyses using only cis-xQTL. MSigDB chose to perform gene target predictions of miRNA by using only miRDB (Chen and Wang, 2020), which might differ from the predictions coming from other methods. The xQTL inference was performed using bulk post-mortem tissue mainly from cortex instead of single cell that might not reflect cell type specific effects. Most xQTL reference data was obtained from European ancestry cohorts, and thus, some of these findings might not be generalizable to non-European GWAS cohorts.

## Data Availability

We used R version 3.6 and Julia version 1.1.1 for the analysis, as they are one of the options available on the cluster. We run the scripts on high performance cluster (Centos 5). Further details can be found in the following link: www.github.com/vipbg/group/wiki/Hardware. We provided analysis scripts for researchers on GitHub page: https://github.com/huseyingedik/MR-GSEA. We also provided the supplementary tables with MR-GSE results as searchable tables on the following link: https://huseyingedik.shinyapps.io/mrgse.

## Author’s contribution

Huseyin Gedik performed the analyses and prepared the data visualization. Silviu-Alin Bacanu supervised the analyses. All authors contributed to the study design and the writing of the manuscript.

## Online resources

MsigDB gene set data (Sep 2022, version 2022.1.Hs*) was obtained from http://www.gsea-msigdb.org/gsea/msigdb/human/collections.jsp GWAS summary statistics for PGC: https://www.med.unc.edu/pgc/download-results/ Blood eQTL (eQTLGen) reference data (.besd, probe, and variant files) : https://molgenis26.gcc.rug.nl/downloads/eqtlgen/cis-eqtl/SMR_formatted/cis-eQTL-SMR_20191212.tar.gz Brain eQTL reference data (.besd, probe, and variant files) : https://yanglab.westlake.edu.cn/data/SMR/BrainMeta_cis_eqtl_summary.tar.gz Blood pQTL (deCODE) reference data: https://download.decode.is/form/folder/proteomics Brain pQTL reference data: https://www.synapse.org/#!Synapse:syn21213340

